# Assessing sustained attention processes and related cerebral activations in typically developing adolescents using the gradual-onset continuous performance task (gradCPT)

**DOI:** 10.1101/2024.10.01.24314449

**Authors:** J. Awada, NB. Fernandez, V. Siffredi, MC. Liverani, J. Miehlbradt, C. Borradori-Tolsa, R. Ha-Vinh Leuchter

**Affiliations:** Division of Development and Growth, Department of Paediatrics, Gynaecology and Obstetrics, Geneva University Hospitals, Geneva, Switzerland; Institute of Bioengineering, Center for Neuroprosthetics, Ecole Polytechnique Fédérale de Lausanne, Switzerland; Department of Radiology and Medical Informatics, Faculty of Medicine, University of Geneva, Switzerland; SensoriMotor, Affective and Social Development Laboratory, Faculty of Psychology and Educational Sciences, University of Geneva, Geneva, Switzerland

## Abstract

**Introduction:** Sustained attention and inhibition processes are fundamental components of attention that mature during adolescence, a transitive period between childhood and adulthood characterized by a rapid behavioral and cognitive development. The current study aimed to provide a better understanding of sustained attention and inhibition processes in typically developing adolescents (n = 26) aged 11-18.

**Methods:** Functional magnetic resonance images (fMRI) were acquired during two different modalities (the *face* and the *scene*) from a previously validated gradual□onset continuous (gradCPT) paradigm to evaluate sustained attention performances. In addition, we performed linear regression analyses to investigate how cerebral activation varied as a function of covariates of interest.

**Results:** We showed a bilateral fronto-parieto-occipito brain activation during response inhibition regardless the type of task. Participants demonstrated better behavioral performances during the *scene* gradCPT. We observed a mainly left-lateralized pattern of activation in a fronto-cingulo-cerebellum area during the *face* gradCPT and an extended bilateral fronto-temporo-parieto-occipital activation during the *scene* gradCPT. Finally, we found associations between brain activity and behavioral attentional responses.

**Conclusion:** This study gives a better understanding of the neural correlates of sustained attention and inhibition in a typically developing adolescent population.

## 1. Introduction

Sustained attention is defined as the capacity to maintain an efficient and consistent level of attention for a long period of time (Langner & Eickhoff, 2012; Posner M, Petersen J., 1990). This capacity is essential in the execution of various daily activities (attending to a lecture or reading a book) and is known to affect academic achievement (Blair & Razza, 2007; Fortenbaugh et al., 2015; Gallen et al., 2023). Another key aspect of attention, closely linked to sustained attention, is inhibition. This involves the ability to suppress dominant or impulsive responses in favor of more appropriate responses (Carlson, 2005; Reck & Hund, 2011). Both sustained attention and inhibition processes appear during childhood and mature during adolescence (Anderson et al., 2001; Best & Miller, 2010; Fortenbaugh et al., 2015; Gallen et al., 2023; Leon-Carrion et al., 2004; Luciana et al., 2005; Luna et al., 2004), that is a critical transitive period between childhood and adulthood marked by a rapid behavioral, socio-emotional, and cognitive development.

Over the past few years, there has been a growing interest in investigating sustained attention performances and their associated neural mechanisms using functional magnetic resonance imaging (fMRI). A common approach to evaluate sustained attention and inhibition is the use of continuous performance tasks (CPTs) (Riccio et al., 2001; Rosvold et al., 1956). Typically, these tasks require participants to continuously assess a constant stream of stimuli with a short interval between them (approximately 1 second)(Rosenberg et al., 2013). Participants are presented with rare target stimuli among frequent non-target stimuli and are asked to respond only to rare target stimuli. In this type of paradigm, participants responses are rare and it is not possible to observe moment-to-moment changes in reaction times (Davies & Parasuraman, 1982; Rosenberg et al., 2013; Szalma et al., 2006; Warm & Jerison, 1984). An alternative approach is the use of modified CPTs, where participants respond to a majority of non-target stimuli while withholding responses to infrequent target stimuli (Conners, 2000; Robertson et al., 1997; Rosenberg et al., 2013). Using this alternative approach, Esterman et al. (2013) introduced a novel not-X CPT called the gradual-onset continuous performance task (gradCPT), whose specificity relies on the implementation of a gradual transition between stimuli, where an image gradually becomes clear while the previous one fades away. This design aims to eliminate “abrupt stimulus onsets” and to increase task demands. Reaction times can therefore be investigated as participants are constantly engaged in responding. Since its introduction, numerous studies employed the gradCPT task to assess sustained attention, in both typical (Esterman et al., 2014, 2017; Fortenbaugh et al., 2018; Kucyi et al., 2016; Rosenberg et al., 2013) and clinical populations (Auerbach et al., 2014; Fortenbaugh, Corbo, et al., 2017; Rosenberg et al., 2016).

Regarding the neural mechanisms underlying attentional processes, recent reviews on healthy adults (Fisher, 2019; Fortenbaugh, DeGutis, et al., 2017; Langner & Eickhoff, 2012) identified typical brain regions activated during different sustained attention tasks, including the prefrontal cortex (medial, bilateral inferior, right midlateral, bilateral pre-supplementary motor area, left dorsal premotor cortex, bilateral ventral premotor cortex), the anterior insula and midcingulate cortex, temporal and parietal areas (right temporo-parietal junction, temporal lobule and intraparietal sulcus), occipital areas (right occipital gyri, left temporo-occipital junction), cerebellum, and subcortical structures (bilateral thalamus). In addition, sustained attention was also studied with fMRI in younger populations such as children and adolescents. In their meta-analysis, Morandini et al. (2020) included 25 fMRI studies using CPTs and go/no-go paradigms and highlighted a sustained attentional network in healthy children and adolescents which is similar to the one previously described in adults population. Although the gradCPT task has demonstrated success in evaluating sustained attention in adults (Esterman et al., 2013, 2016; Jun & Lee, 2021) and in populations of children and adolescents at risk (Auerbach et al., 2014; Rosenberg et al., 2016), it has not been used to investigate the brain correlates of sustained attention and inhibition using this task in typically developing adolescents.

Therefore, the present study aimed to provide a better understanding of sustained attention and inhibition processes in adolescents using a validated fMRI gradCPT paradigm comprising two distinct modalities (Rosenberg et al., 2016). We investigated both behavioral performance and brain activation patterns in a cohort of typically developing adolescents and examined how executive and attentional abilities relate to brain activity involved in sustained attention.

## 2. Methods

### 2.1 Participants

Twenty-nine adolescents from 11 to 18 years old (M = 15 years, SD = 2) were recruited through community and were born after 37 gestational weeks, developed typically, and attended mainstream schools. Participants were excluded if they had an intelligence quotient below 70, any sensory or physical disabilities (such as blindness, hearing loss, cerebral palsy), or an insufficient understanding of French or English. To estimate general intellectual functioning, we used the General Ability Index (GAI) from the Wechsler Intelligence Scale for Children - 5th Edition (WISC-V) (Wechsler, 2014). The GAI has a mean of 100 and a standard deviation of 15. The Largo score was used to estimate socio-economic status based on maternal education and paternal occupation (Largo et al., 1989). Higher socio-economic scores indicate lower socio-economic status levels.

The current study was approved by the Swiss Ethics Committee on research involving humans (ID: 2015-00175). All participants and their principal caregiver gave written and informed consent in accordance with regulation of the ethics committee at the University Hospital of Geneva. A gift voucher worth 50 Swiss francs was given to all participants as a token of appreciation for their involvement in the study.

### 2.2 Executive and attentional behavioral measures

#### 2.2.1 Conners’s Rating scale third edition (Conners 3-SR)

The *Conners self-report questionnaire* was used to assess adolescents’ attentional abilities in their daily lives and allows the assessment of symptoms associated with attention disorders with or without hyperactivity. It consists of 97 questions, and categorizing the items results in two subscales: the inattention scale and the hyperactivity/impulsivity scale. The results are presented as T-scores.

#### 2.2.2 Behavior Rating Inventory of Executive Functioning (BRIEF)

Parents completed the *BRIEF questionnaire*, which provides an assessment of executive functions within home environment for children and adolescents aged between 5 and 18 years old (Gioia et al., 2014). Commonly used for clinical assessment, it includes 8 clinical scales of executive functions (emotional control, inhibition, flexibility, initiation, material organization, working memory, planning/organization, and control) that can be categorized into two standardized subscales. The *Behavioral Regulation Index (BRI)* combines inhibit, shift, and emotional control scales, while the *Metacognition Index (MI)* with initiate, organization of materials, working memory, plan/organize and monitor scales. The *Global Executive Composite index (GEC)* is obtained with the sum of scores from all 8 scales. In the present study, we used T-scores of the 3 indexes (BRI, MI, and GEC) to assess the executive abilities of our typically developing adolescents.

#### 2.2.3 Test of Attentional Performance (TAP)

Computerized tasks of the Test of Attentional Performance (TAP) (Zimmermann & Fimm, 2002) were used to test attentional skills and all instructions were given orally by the experimenter. To assess inhibitory control, we chose two *Go/No-Go* paradigms (1 target against 3 stimuli and 2 targets against 5 stimuli) and analyzed the number of errors that is an interesting indicator for impulsivity and control deficit (Zimmermann & Fimm, 2002, p 51).

### 2.3 Experimental fMRI paradigm: Gradual-Onset Continuous Performance task

Stimuli presentation and response recording through MRI-compatible response buttons were controlled with a Psychophysics Toolbox extension (https://psychtoolbox.org) implemented in Matlab 2019b (Mathworks Inc., Natick, MA, USA).

The experimental paradigm used in the current study encompasses two distinct versions of the gradual-onset continuous performance task (gradCPT), each representing distinct modalities (*face* and *scene)* which have been previously validated and described by Rosenberg and colleagues (Rosenberg et al., 2013; Rosenberg et al., 2016). In the *face* modality, female faces were considered as target trials and male faces as non-target trials, while in the *scene* modality, mountain images were considered as target trials and city images as non-target trials (see *Figure 1*).

**Figure 1:**
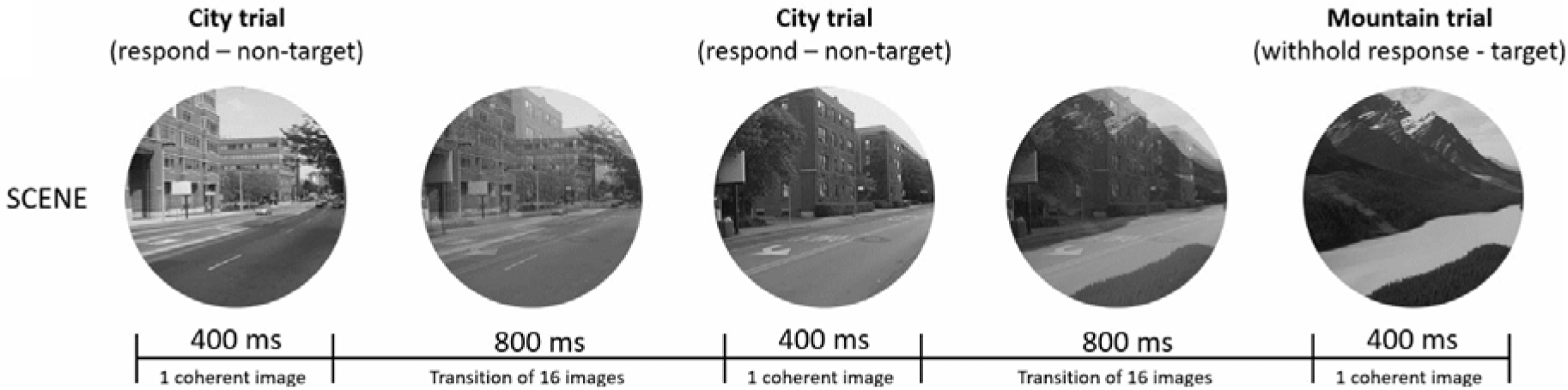
GradCPT. Illustration of the gradCPT paradigm, for scene modality. Target trials (mountains) are presented 10% of the time and must be withheld, while non target trials (city scenes) are presented 90% of the time and must be answered by pressing a button. For each trial (1’200ms duration), stimuli gradually transition from the current to the next trial during 800ms (succession of 16 images in total), followed by a 400ms pause during which the stimulus is fully coherent (Rosenberg et al., 2013).

These two versions of the gradCPT task had a duration of six minutes each and visual stimuli gradually transitioned between the target and non-target stimuli at a constant rate. In each trial (1’200 milliseconds (ms) duration), the image transition took 800ms, followed by a 400ms pause during which the stimulus fully cohered. For each modality, visual stimuli were randomly presented, with target trials appearing 10% of the time (60 trials) while non-target trials occurring 90% of the time (540 trials). There was no repetition of the ongoing stimulus in the upcoming trial. Participants were asked to respond as fast and as accurately as possible to non-target stimuli (male faces or city) by pressing the response button and to withhold response to target stimuli (female faces or mountains). See *Supplemental material* for more details about accurate non-target and target trials (Rosenberg et al., 2013).

### 2.4 Behavioral data analyses

Sustained attentional performance was assessed using the percentage of correct answers (accuracy, AC), sensitivity scores (*d’*), and the reaction times coefficient of variation (RT_CV_). All statistical analyses were performed using RStudio version 4.2.1 (R Core Team, 2022) and Statistica software version 14.0.0 (*TIBCO Statistica*, 2020).

The percentage of correct answers (AC) of non-target and target trials was calculated for each participant and each modality separately. Sustained attentional performances were analyzed by entering AC scores in a Mann-Whitney test with modality (*face* and *scene*) and stimuli (non-target and target) as within-subject levels.

The *sensitivity* score (*d’*) is a measure of performance and considers the proportion of correct non-target (*HIT rate*) and the proportion of incorrect target (*FALSE ALARM rate*). It was calculated for each participant and each modality separately using the following formula: *d’= z(HIT rate) - z(FALSE ALARM rate)*(for more details, see Rosenberg et al., 2016). We compared sustained attention performances between *face* and *scene* modalities with *d’* scores using a simple t-test.

Previous studies have found that reaction times (RT) variability is a significant indicator of attentional state and a unique component of the performance of sustained attention (Rosenberg et al., 2013; Rosenberg et al., 2015). The *reaction times coefficient of variation* (RT_CV_) was calculated by dividing the standard deviation of reaction times (RTs) by the mean RTs (Fortenbaugh et al., 2018; Rosenberg et al., 2015; Urbain et al., 2019). We calculated RT_CV_ for correct non-target stimuli for each participant and for each modality independently. Differences between modalities on RT_CV_ were analyzed using simple t-tests.

Additionally, we assessed the effect of age, socio-economic status (Largo scale), executive functions (BRIEF scores) and attentional abilities (Conners 3-SR and TAP scores) on sustained attentional performances. We conducted two distinct multiple linear regressions, incorporating *d’* and RT_CV_ scores as the dependent variables, Modality (*face* vs *scene*) as a within subject factor, and a covariate as a single factor.

### 2.5 fMRI analyses

#### 2.5.1 Functional Magnetic Resonance Imaging (MRI) acquisition

MRI data were acquired at the Campus Biotech (Geneva, Switzerland) using a Siemens 3T Magnetom Prisma scanner and a standard 64-channel head coil. Structural T1-weighted magnetization-prepared rapid gradient-echo (MP-RAGE) sequences were acquired using the following parameters: voxel size = 0.9 × 0.9 × 0.9 mm, TR/TE = 2300/ 2.32 ms, flip angle = 8°, and FOV = 240 x 240 mm. Functional T2*-weighted images were acquired using multi-slice gradient-echo-planar imaging (EPI) sequences and the following parameters: voxel size = 2 x 2 x 2 mm, TR/TE = 720/33ms, flip angle = 52°, and FOV = 208 x 208 mm. Finally, a fieldmap was acquired for each participant, with TR/TE_1_/TE_2_ = 627/5.19/7.65 ms, flip angle = 60°, voxel size = 2 x 2 x 2 mm, and FOV = 208 x 208 mm.

#### 2.5.2 MRI data preprocessing

Statistical Parametric Mapping software (SPM12; Wellcome Department of Imaging Neuroscience, UCL, UK) implemented in MATLAB R2019a (The MathWorks, Inc., Natick, Massachusetts, United States) was used for the preprocessing and analysis of MRI images.

Following an inhouse preprocessing pipeline (Freitas et al., 2021; Liverani et al., 2020), fMRI images were spatially realigned and unwarped to correct for motion artefacts and geometric distortions (Hutton et al., 2002) for each participant. Functional images were then co-registered with the structural images in the individual’s own brain space. A segmentation step using the SPM12 segmentation algorithm was performed on structural images to identify distinct tissue probability maps, i.e., grey matter, white matter, and cerebrospinal fluid (Ashburner & Friston, 2005) and to generate a study specific template (using Diffeomorphic Anatomical Registration using Exponential Lie algebra) (DARTEL, Ashburner, 2007). Eventually, functional images were normalized to the DARTEL template and smoothed with a 6 mm isotropic Gaussian kernel. As previously applied by researchers in our lab (see Freitas et al., 2021; Liverani et al., 2020), we finally assessed head motion based on Framewise Displacement (FD, see Power et al., 2012), which was calculated for each modality (*face* or *scene*) separately and used as an exclusion criterion. Only one participant was excluded from the current study as more than 20% of his frames showed excessive motion (frames with FD greater than 0.5mm, including one frame before and two after those).

#### 2.5.3 fMRI analysis

Individual first level analyses were performed with a General Linear Model (GLM). The model includes the two modalities with target (T) and non-target (NT) conditions. Additionally, the six realignment parameters (head motions) were entered as non-interest covariates. Trial onsets were aligned with the beginning of image changes, lasting for 0.57 seconds, and adjusted using the canonical hemodynamic response function (cHRF). For each participant, first-level T-contrasts were determined by comparing the two experimental conditions (T > NT) and the two experimental modalities (*face* > *scene* or *scene* > *face*).

Resulting first-level statistical parametric maps were entered into a second-level (whole-brain) flexible factorial model comprising modalities (*face* and *scene*), stimuli (T and NT), and subject as separate factors. First, the main effect of task was measured by comparing the two modalities (*face* > *scene* or *scene* > *face*). Second, the main effect of response inhibition was measured by comparing the target to non-target conditions (T > NT). Then, the effect of response inhibition on modality was measured by contrasting modalities and stimuli (*face* (T > NT) > *scene* (T > NT) or *scene* (T > NT) > *face* (T > NT)). The direction of the interaction was confirmed by applying the effect of response inhibition for a specific task (*face* (T > NT) or *scene* (T > NT) respectively) as an inclusive mask to restrict the direction of the contrast.

Moreover, we investigated how cerebral activity during response inhibition varied as a function of covariates of interest (*d’* scores and RT_CV_, age at testing, socio economic status, as well as BRIEF, Conners 3-SR, and TAP scores) by performing distinct univariate linear regression analyses. We extracted individual beta values associated with significant peak activations from the univariate linear regression analyses. This was achieved by employing spheres (27 voxels) centered on the main peak coordinates identified by covariates SPM analyses. We then conducted correlations analyses between beta values and corresponding covariates of interest using R Studio.

For second-level whole-brain analyses, we reported p-values with a statistical threshold of p_unc_ < 0.001 uncorrected for multiple comparisons, with a minimum size of 50 voxels (Lieberman & Cunningham, 2009). All findings are presented in MNI space.

## 3 Results

### 3.1 Participants characteristics

Of the 29 adolescents enrolled, one participant was excluded as he did not complete the scanner session entirely, one participant was excluded due to excessive motion, and one participant was excluded because he never withheld response during the task. The final sample included 26 participants. Participants were balanced between male (n=13, 50%), and female (n=13, 50%). Participants presented a mean general ability index of 110.36 (SD = 11.11) and a mean socio-economic status (Largo score) of 3.00 (SD = 1.57).

### 3.2 Executive and attentional behavioral results

Executive and attentional neurobehavioral scores are shown in *Table 1* and are represented in T-scores. Overall, all executive and attentional behavioral scores were within the average range.

**Table 1:** A) Executive and attentional behavioural results. Scores resulting from the parent-reported BRIEF questionnaire, the self-reported Conners 3-SR and the TAP are reported in T scores. B) GradCPT behavioural performances for the Face and the Scene tasks. Mean accuracy (a), mean sensitivity scores (b), and reaction times variability (c) are presented with standard deviation in brackets. Statistical comparisons between the two tasks were performed using Wilcoxon signed rank test (W) for accuracy values, and a paired-sample Student t-test (t) for sensitivity scores and reaction times variability.

| A) Executive and attentional neurobehavioural results |  | Mean T scores (SD) |  |
| --- | --- | --- | --- |
| Parent-reported BRIEF questionnaire |  |  |  |
| Global Executive Composite Index (GEC) (T) |  | 52.19 (10.9) |  |
| Metacognition Index (MI) (T) |  | 52.27 (10.9) |  |
| Behavioral Regulation Index (BRI) (T) |  | 51.08 (12.2) |  |
| Conners 3-SR |  |  |  |
| Inattention (T) |  | 54.81 (10.3) |  |
| Hyperactivity (T) |  | 56.77 (11.6) |  |
| Neuropsychological testing – TAP |  |  |  |
| Go/No-Go (2 stimuli, 1 target) (Nb of errors (T)) |  | 42.42 (7.1) |  |
| Go/No-Go (5 stimuli, 2 targets) (Nb of errors (T)) |  | 46.77 (7.5) |  |
| B) GradCPT Behavioural performances | Face task | Scene task | Task comparison |
| (a) Accuracy |  |  |  |
| Percentage of Correct Non-Target | 86.5% (14%) | 93.9 (7%) | W = 246.5, p = .096 |
| Percentage of Correct Target | 22.1% (17%) | 28.1 (22%) | W = 288.5, p = .369 |
| (b) Sensitivity Scores (d') |  |  |  |
| d' = z(HIT rate) – z(FALSE ALARM rate) | 0.44 (0.43) | 1.12 (0.59) | t(25) = -5.11, p <.001 |
| (c) Reaction times variability (RT<sub>CV</sub>) |  |  |  |
| RT<sub>CV</sub> = standard deviation of RT/mean RT | 0.29 (0.04) | 0.25 (0.04) | t(25) = 4.99, p <.001 |

### 3.3 GradCPT behavioral results

Detailed gradCPT behavioral results (AC, *d’*, and RT_CV_) are presented in *Table 1*. Analyses of sustained attentional performances revealed a main effect of the modality with better *d’* scores (p < .001) and RT_CV_ (p < .001) in the *scene* compared to the *face* gradPCT. Accuracy analyses showed similar percentages of correct responses between the *face* and the *scene* gradCPT, for both non-target (W = 246.5, p = .096) and target stimuli (W = 288.5, p = .369). For both modalities, the percentage of correct responses to non-target stimuli was significantly higher compared to target stimuli (p < .001).

Additionally, the multiple linear regression using *d’* scores did not reveal a significant effect of any covariate of interest. However, the multiple linear regression using RT_CV_ scores revealed that the inattention scores from Conners 3-SR questionnaire [t(38) = 2.05, p = .047] and number of errors during the Go/No-Go computerized task (2 stimuli, 1 target) from the TAP [t(38) = -3.13, p = .003] explained 49.9% of the variation in the RT_CV_ scores. These results are shown in *Table 2*.

**Table 2:**
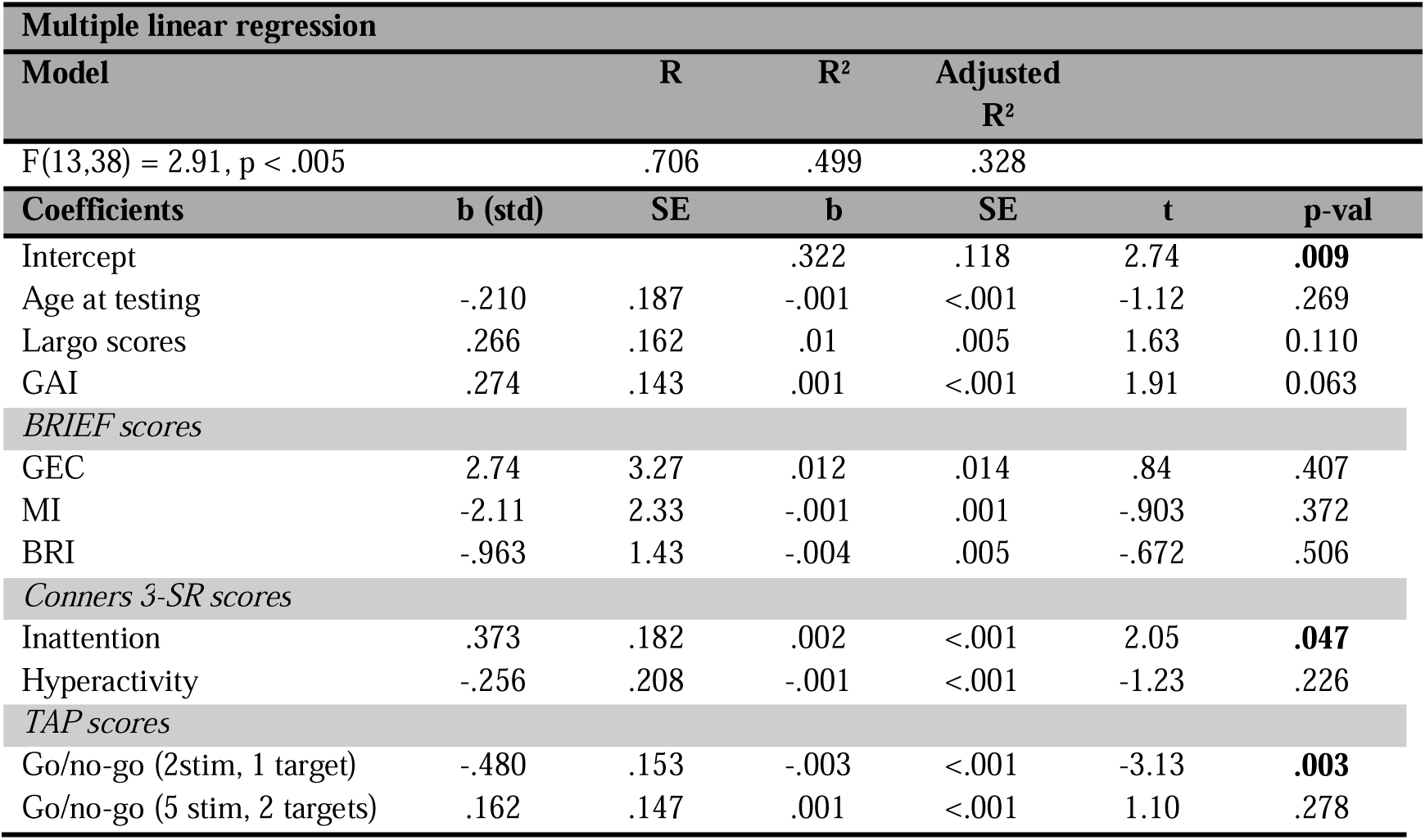
Analyses of GradCPT behavioral performances including covariates. Statistical comparisons between the two tasks were performed using two multiple linear regressions, incorporating RT_CV_ scores as the dependant variable, Task (Face vs Scene) as a within subject factor, and a covariate (i.e., either age, socio-economic status, general ability index (GAI), BRIEF (GEC, MI, and BRI), Conners 3-SR, and TAP scores) as a single factor.

### 3.4 fMRI results – Whole-brain activations during the gradCPT paradigm

The main effect of response inhibition regardless the type of task revealed a bilateral fronto-parieto-occipito pattern of brain activity (see *Table 3, Figure 2*). This pattern includes frontal regions (frontal operculum, the inferior, superior, and middle frontal gyrus (IFG, SFG, and MFG, respectively), precentral gyrus, motor areas), parietal (both the inferior and superior parietal lobule (the IPL and the SPL, respectively) and the supramarginal gyrus (SMG), occipital (the fusiform gyrus and the middle occipital gyrus (MOG)), and other regions such as the anterior cingulate cortex and the midcingulate cortex (aMCC), the anterior insula, the cerebellum, and the thalamus.

**Figure 2:**
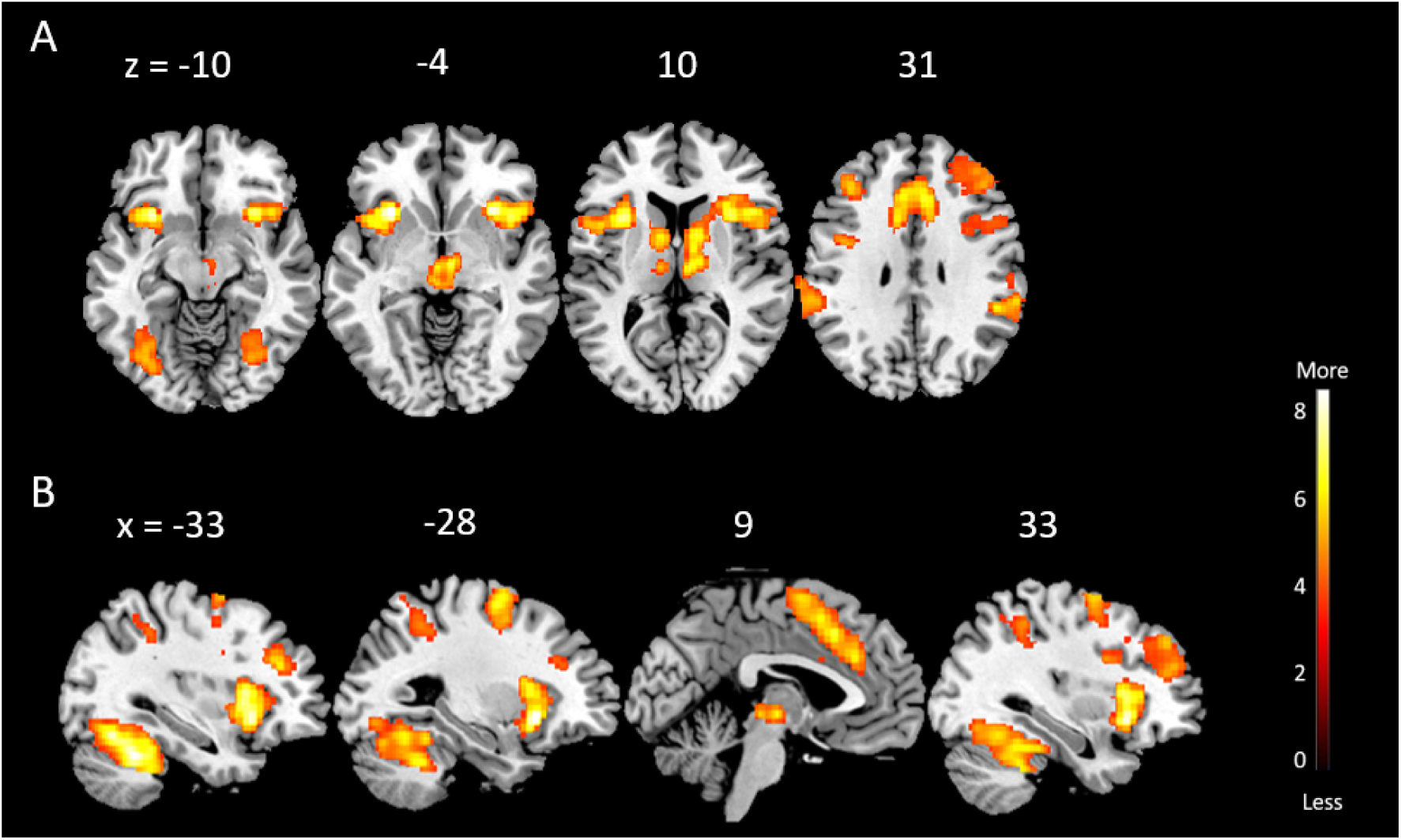
Cerebral areas engaged during response inhibition trials (Target > Non-Target). All clusters are significant at peak-level at p < 0.001, uncorrected for multiple comparisons, with a minimum size of 50 voxels. Activations are displayed on a template image and numbers indicate z (A. axial view) and x (B. sagittal view) coordinates in a MNI space.

**Table 3:**
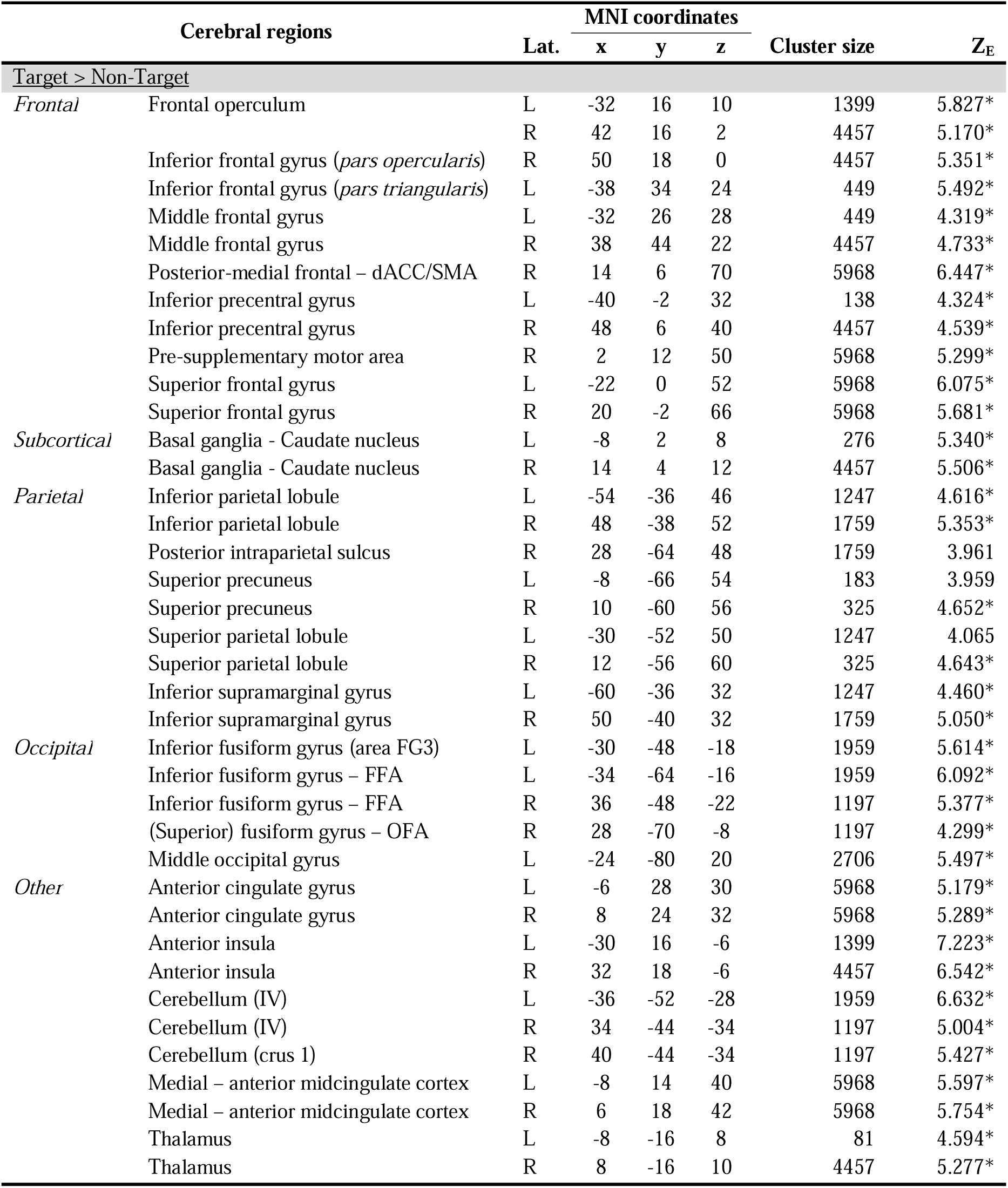
Regional cerebral activations during response inhibition regardless the type of task (Target > Non-Target). All reported clusters are significant at peak-level at p < 0.001, uncorrected for multiple comparisons, with a minimum size of 50 voxels. Peaks that are significant after correction for multiple comparison are indicated with an asterisk (*FDR-corrected). Abbreviation: Lat.: Hemisphere lateralisation (L = left ; R = right), Z-score (Z_E_) values refer to the activation maxima to the SPM coordinates.

The same contrast performed for the *face* gradCPT (see *Table 4, Figure 3*) showed a mainly left-lateralized pattern of activation that includes frontal regions (the precentral gyrus, the supplementary motor area (SMA), and the SFG), the aMCC, as well as the posterior cerebellum. The response inhibition related to the *scene* gradCPT (see *Table 4, Figure 4*) revealed an extended bilateral brain activity located in frontal areas, including frontal gyri (inferior, middle, and superior), motor area, precentral gyrus, as well as bilateral temporal activations in both the inferior and middle temporal gyri (ITG and MTG, respectively). In addition, parietal regions such as the intraparietal sulcus, both the IPL and SPL, precuneus, and the SMG, as well as occipital regions including inferior occipital gyrus (IOG), the MOG, and fusiform gyrus were also more activated in the *scene* compared to the *face* gradCPT. Analyses also revealed brain activity in the anterior cingulate cortex, the anterior insula, the cerebellum, and the thalamus.

**Figure 3:**
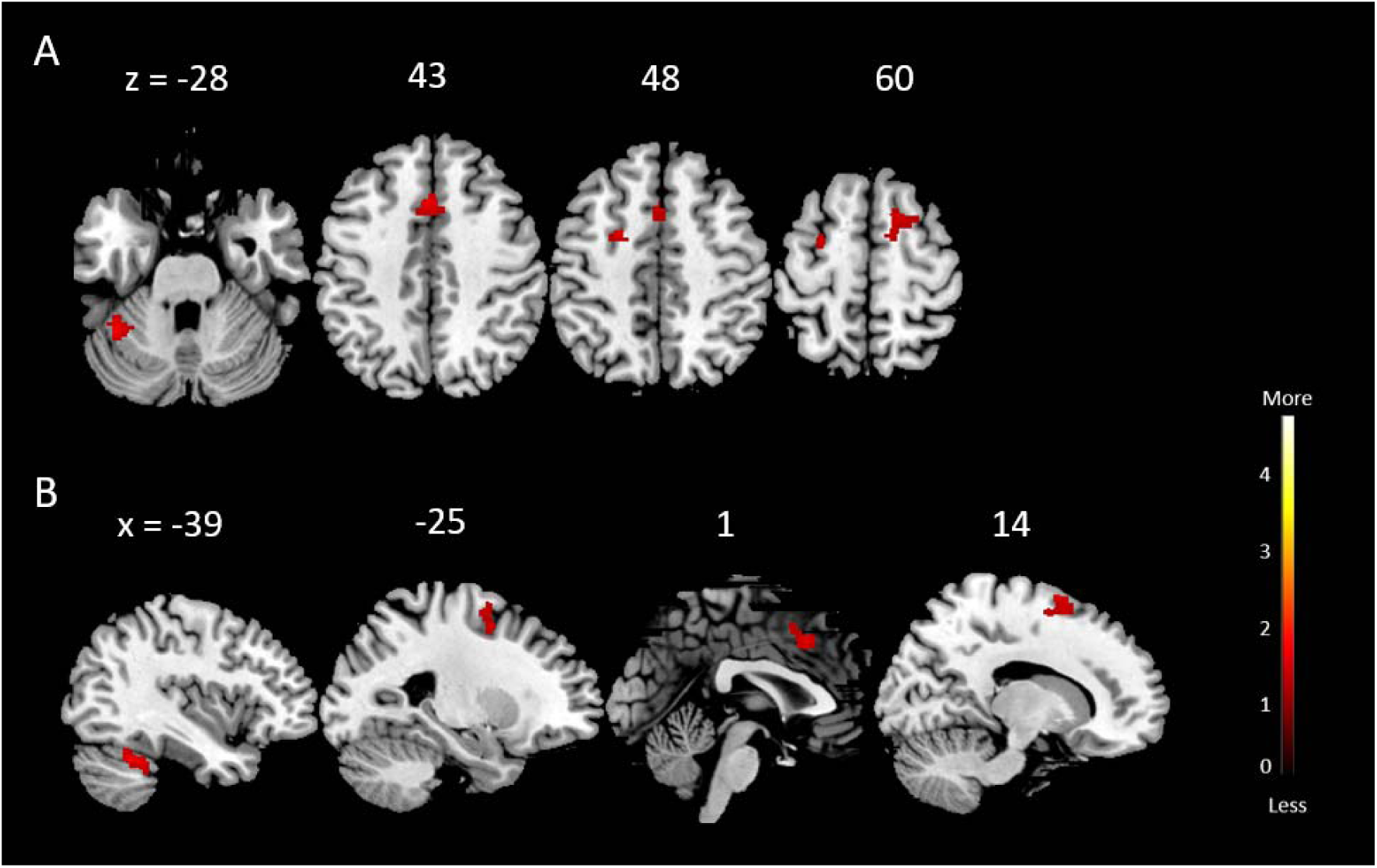
Cerebral areas engaged during response inhibition trials in the Face task (Target > Non-Target). All clusters are significant at peak-level at p < 0.001, uncorrected for multiple comparisons, with a minimum size of 50 voxels. Activations are displayed on a template image and numbers indicate z (A. axial view) and x (B. sagittal view) coordinates in a MNI space.

**Figure 4:**
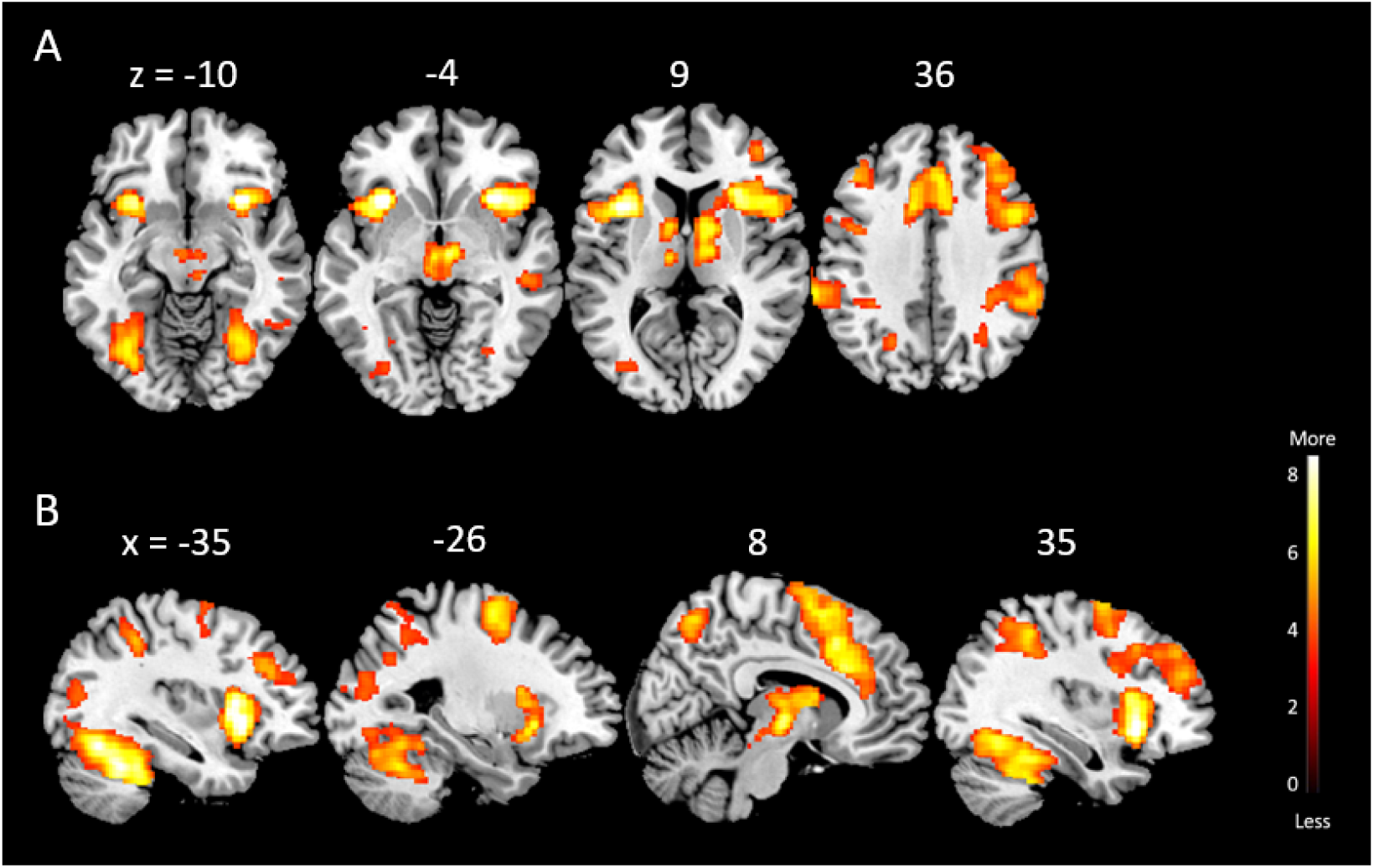
Cerebral areas engaged during response inhibition trials in the Scene task (Target > Non-Target). All clusters are significant at peak-level at p < 0.001, uncorrected for multiple comparisons, with a minimum size of 50 voxels. Activations are displayed on a template image and numbers indicate z (A. axial view) and x (B. sagittal view) coordinates in a MNI space.

**Table 4:** Regional cerebral activations associated with performance of a response inhibition during the Face and Scene tasks. Clusters are significant at peak-level at p < 0.001, uncorrected for multiple comparisons, with a minimum size of 50 voxels. Clusters that are significant after correction for multiple comparison are indicated with an asterisk (*FDR-corrected). Abbreviation: Lat.: Hemisphere lateralisation (L = left ; R = right), Z-score (Z_E_) values refer to the activation maxima to the SPM coordinates.

| Cerebral regions |  |  | MNI coordinates |  |  | Cluster size | Z<sub>E</sub> |
| --- | --- | --- | --- | --- | --- | --- | --- |
|  |  |  | x | y | z |  |  |
| Face (Target>Non-Target) |  |  |  |  |  |  |  |
| Frontal | Superior – precentral gyrus | L | -26 | -2 | 60 | 118 | 3.394 |
|  | Posterior-medial frontal/SMA | L | 0 | 18 | 44 | 155 | 3.995 |
|  | Superior frontal gyrus | L | -22 | 0 | 52 | 118 | 4.267 |
|  | Superior frontal gyrus | R | 16 | 10 | 62 | 183 | 3.761 |
| Other | Cerebellum (VI) | L | -40 | -46 | -30 | 112 | 4.254 |
|  | Medial – anterior midcingulate gyrus | L | -8 | 14 | 38 | 155 | 3.225 |
| Scene (Target>Non-Target) |  |  |  |  |  |  |  |
| Frontal | Inferior frontal gyrus | L | -52 | 10 | 12 | 1499 | 4.695* |
|  | (p. Opercularis) | R | 50 | 18 | 0 | 12784 | 5.886* |
|  | Middle frontal gyrus | L | -40 | 36 | 26 | 524 | 5.230* |
|  | Superior frontal gyrus | L | -20 | -2 | 62 | 12784 | 5.733* |
|  | Superior frontal gyrus | R | 18 | 0 | 60 | 12784 | 5.725* |
|  | Superior – precentral gyrus | L | -60 | 8 | 30 | 1499 | 3.639 |
|  | Posterior-medial – SMA | R | 14 | 4 | 70 | 12794 | 6.414* |
|  | Medial – pre-SMA | L | -10 | 12 | 42 | 12784 | 5.823* |
| Temporal | Inferior temporal gyrus | R | 50 | -56 | -14 | 1796 | 3.947 |
|  | Middle temporal gyrus | R | 50 | -28 | -4 | 96 | 4.083 |
| Parietal | Anterior – intraparietal sulcus | L | -24 | -58 | 46 | 1906 | 3.761 |
|  | Inferior parietal lobule | L | -40 | -44 | 46 | 1906 | 4.908* |
|  | Inferior parietal lobule | R | 48 | -38 | 52 | 2875 | 6.337* |
|  | Superior parietal lobule | L | -30 | -50 | 48 | 1906 | 4.144 |
|  | Superior parietal lobule | R | 28 | -64 | 48 | 2875 | 4.768* |
|  | Superior – precuneus | R | 10 | -64 | 56 | 381 | 5.058* |
|  | Superior – supramarginal gyrus | L | -62 | -48 | 42 | 1906 | 3.672 |
|  | Superior – supramarginal gyrus | R | 62 | -38 | 30 | 2875 | 5.363* |
| Occipital | Inferior occipital gyrus | L | -33 | -84 | 0 | 2863 | 3.363 |
|  | Middle occipital gyrus | L | -30 | -78 | 22 | 2863 | 4.219* |
|  | Superior – fusiform gyrus | L | -30 | -48 | -18 | 2863 | 6.070* |
|  | Inferior – fusiform gyrus (FFA) | L | -34 | -64 | -16 | 2863 | 6.610* |
|  | Inferior – fusiform gyrus (FFA) | R | 34 | -62 | -14 | 1796 | 5.613* |
| Other | Medial – anterior cingulate cortex | R | 8 | 24 | 28 | 12794 | 5.790* |
|  | Lateral – anterior cingulate cortex | R | 8 | 18 | 42 | 12794 | 5.933* |
|  | Anterior insula | L | -36 | 12 | 2 | 1499 | 7.235* |
|  | Anterior insula | R | 32 | 18 | -8 | 12794 | 7.586* |
|  | Cerebellum (VI) | L | -32 | -54 | -32 | 2863 | 6.138* |
|  | Cerebellum (VI) | R | 34 | -54 | -30 | 1796 | 5.264* |
|  | Cerebellum (crus 1) | L | -40 | -56 | -28 | 2863 | 6.748* |
|  | Cerebellum (crus 1) | R | 40 | -44 | -34 | 1796 | 5.682* |
|  | Thalamus | R | 6 | -12 | -2 | 12794 | 5.985* |

### 3.5 Associations between brain activation related to response inhibition and behavioral measures

Multiple linear regressions including covariates were performed for each modality separately to assess how individual differences in demographic characteristics (age at testing, socio-economic status) as well as executive and attentional abilities (*d’*, RT_CV_, well as BRIEF, Conners 3-SR scores, and TAP scores) might influence the response inhibition brain activity.

Regarding the *face* gradCPT, we found positive associations between RT_CV_ (higher reaction time variability) and brain activation in frontal regions, including bilateral superior frontal gyrus and dACC/SMA, in parietal regions such as bilateral angular gyrus, as well as in temporal regions, in the left MTG and the left STG. These results are showed in *Table 5 (*see *Table SM3* in *Supplemental material* for Pearson’s correlation coefficients). We did not find any association between *d’* scores and response inhibition-induced brain activity. There were also positive associations between the Conners’ inattention scores and brain activation in regions of frontal (the superior precentral gyrus), temporal (the STG), parietal (the superior precuneus and the SPL), occipital (the calcarine sulcus, the superior cuneus, the lingual gyrus, the superior occipital gyrus (SOG)), and insula lobes (see *Table 6*) (see *Table SM4* for Pearson’s correlation coefficients). Additionally, we found positive correlations with the number of errors in the Go/No-Go task (2 stimuli – 1 target) in parietal (the inferior precuneus), occipital (the inferior cuneus) and subcortical brain regions (the putamen). One positive correlation with the number of errors in the Go/No-Go task (5 stimuli – 2 targets) was found in the superior precentral gyrus. These results are shown on *Table 7* (see *Table SM5* for Pearson’s correlation).

**Table 5:**
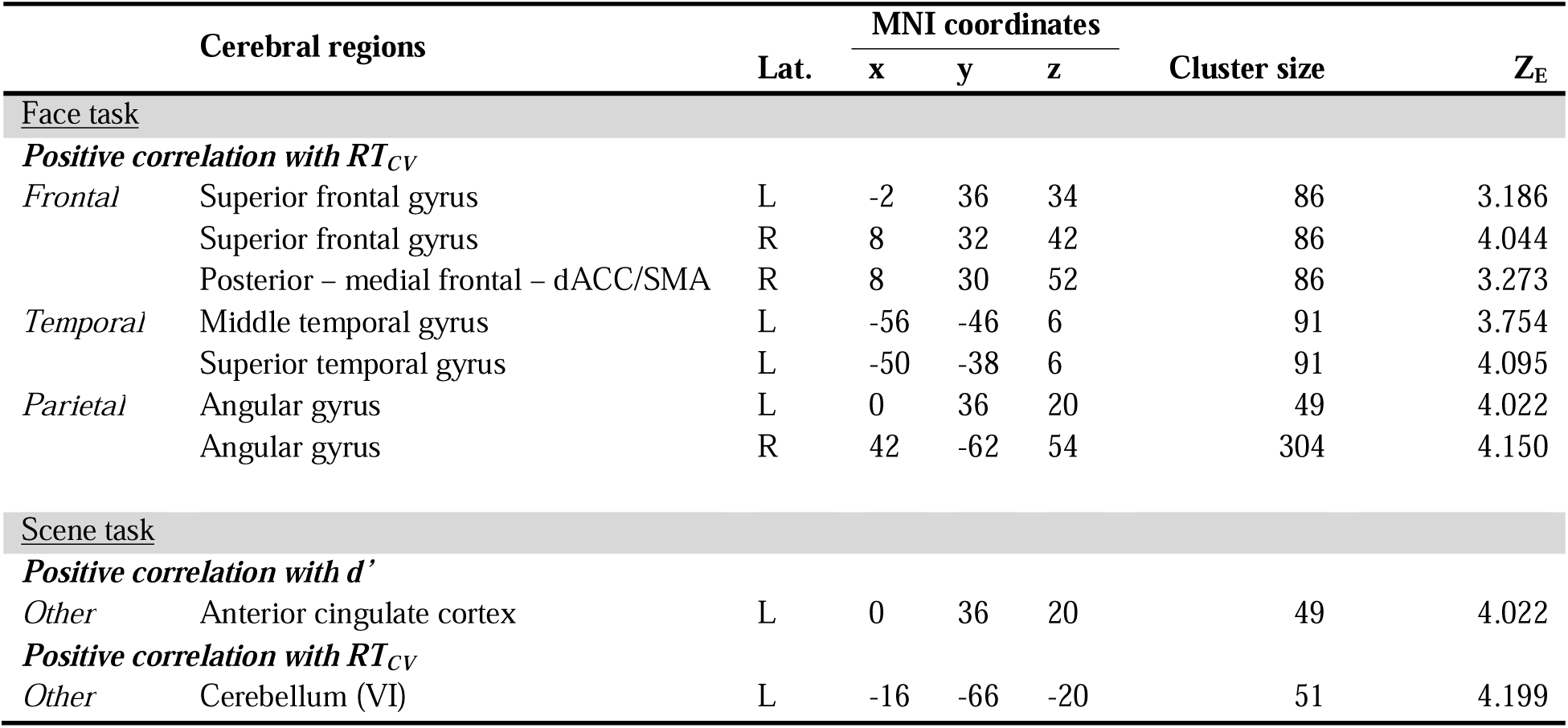
Regional cerebral activations associated with behavioural measures (d’ and RT_CV_) during the Scene task during response inhibition (Target>Non-Target). Clusters are significant at peak-level at p < 0.001, uncorrected for multiple comparisons, with a minimum size of 50 voxels. Abbreviation: Lat.: Hemisphere lateralisation (L = left), Z-score (Z_E_) values refer to the activation maxima to the SPM coordinates.

**Table 6:**
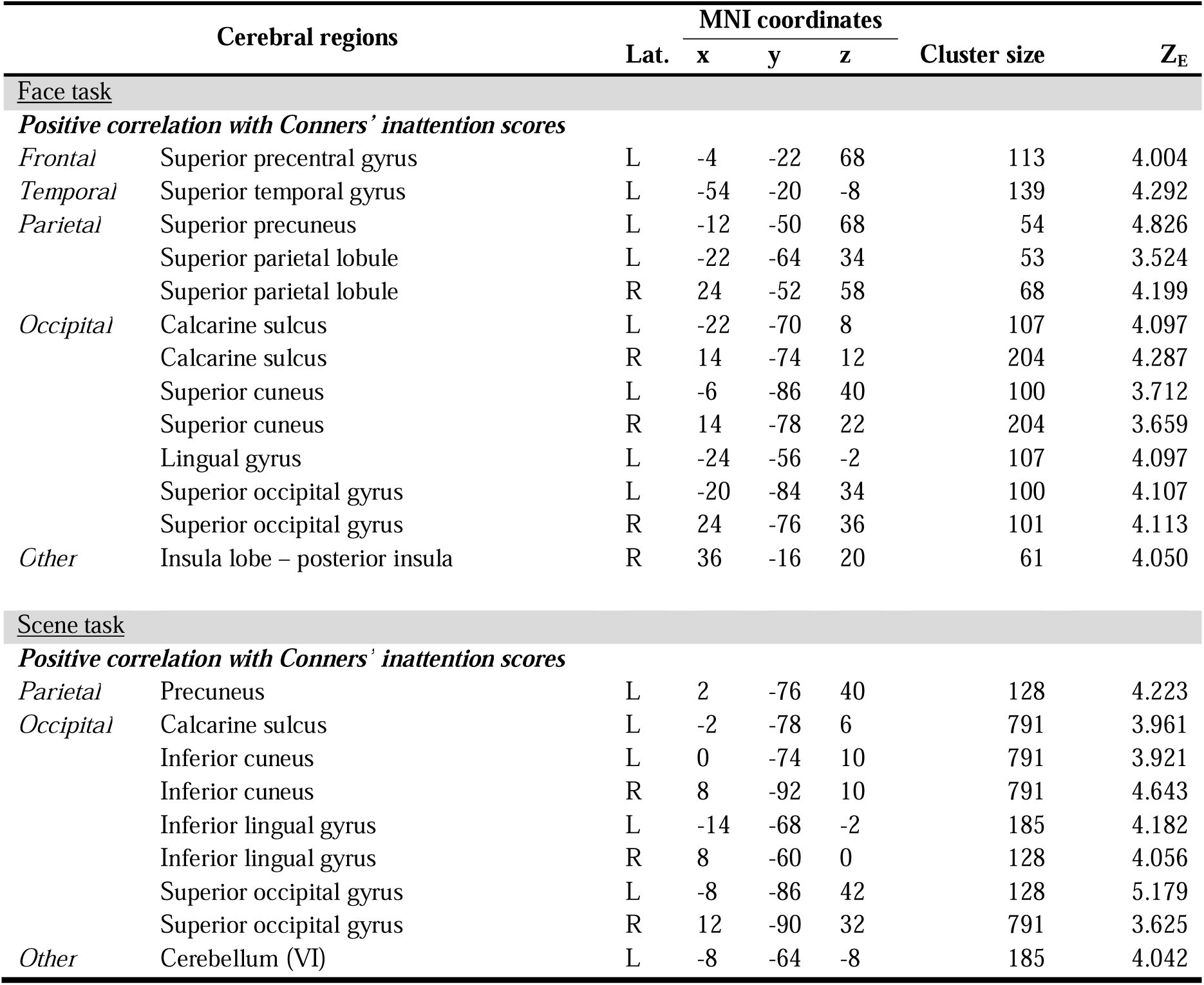
Regional cerebral activations associated with Conners’ inattention scores during the Face and Scene tasks during response inhibition (Target>Non-Target). Clusters are significant at peak-level at p < 0.001, uncorrected for multiple comparisons, with a minimum size of 50 voxels. Abbreviation: Lat.: Hemisphere lateralisation (L = left), Z-score (Z_E_) values refer to the activation maxima to the SPM coordinates.

**Table 7:**
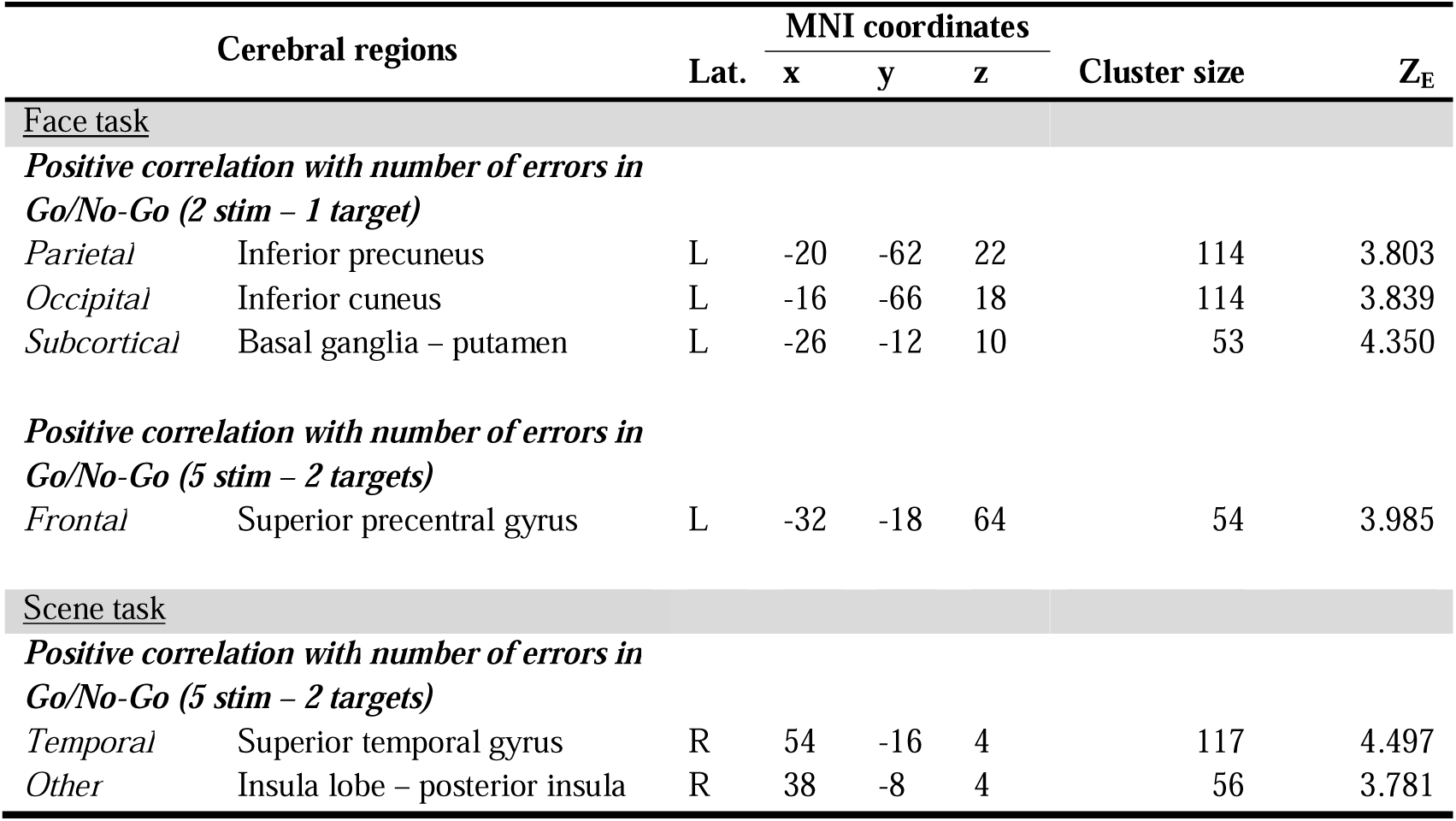
Regional cerebral activations associated with number of errors in the Go/No-Go task of the TAP during the Face and Scene tasks during response inhibition (Target>Non-Target). There are two types of Go/No-Go task: one with 2 presented stimuli and 1 target (Go/No-Go (2 stim – 1 target)) and one with 5 presented stimuli and 2 targets (Go/No-Go (5 stim – 2 targets)). Clusters are significant at peak-level at p < 0.001, uncorrected for multiple comparisons, with a minimum size of 50 voxels. Abbreviation: Lat.: Hemisphere lateralisation (L = left), Z-score (Z_E_) values refer to the activation maxima to the SPM coordinates.

| Cerebral regions |  | Lat. | MNI coordinates |  |  | Cluster size | Z<sub>E</sub> |
| --- | --- | --- | --- | --- | --- | --- | --- |
|  |  |  | x | y | z |  |  |
| <u>Face task</u> |  |  |  |  |  |  |  |
| **Positive correlation with number of errors in** |  |  |  |  |  |  |  |
| **Go/No-Go (2 stim – 1 target)** |  |  |  |  |  |  |  |
| *Parietal* | Inferior precuneus | L | -20 | -62 | 22 | 114 | 3.803 |
| *Occipital* | Inferior cuneus | L | -16 | -66 | 18 | 114 | 3.839 |
| *Subcortical* | Basal ganglia – putamen | L | -26 | -12 | 10 | 53 | 4.350 |
| **Positive correlation with number of errors in** |  |  |  |  |  |  |  |
| **Go/No-Go (5 stim – 2 targets)** |  |  |  |  |  |  |  |
| *Frontal* | Superior precentral gyrus | L | -32 | -18 | 64 | 54 | 3.985 |
| <u>Scene task</u> |  |  |  |  |  |  |  |
| **Positive correlation with number of errors in** |  |  |  |  |  |  |  |
| **Go/No-Go (5 stim – 2 targets)** |  |  |  |  |  |  |  |
| *Temporal* | Superior temporal gyrus | R | 54 | -16 | 4 | 117 | 4.497 |
| *Other* | Insula lobe – posterior insula | R | 38 | -8 | 4 | 56 | 3.781 |

Regarding the *scene* gradCPT, we found positive associations between gradCPT behavioral performances and brain activations. Specifically, participants with higher *d’* scores (better gradCPT attentional performances) activated more the anterior cingulate (see *Table 5, Figure 5*), while participants with higher RT_CV_ exhibited larger brain activation in the left posterior cerebellum (see *Table 5*) (see *Table SM3* for Pearson’s correlation coefficients). Regarding attentional abilities, positive associations between Conners’ inattention scores and brain activation were found in parietal regions including the precuneus, as well as occipital regions including the calcarine sulcus, the inferior cuneus, the inferior lingual gyrus and the SOG, and the posterior cerebellum (see *Table 6*) (see *Table SM4* for Pearson’s correlation coefficients). Finally, we found that activation in the STG, as well as the insula lobe were positively correlated with the number of errors in Go/No-Go task (5 stimuli – 2 targets) (see *Table 7*) (see *Table SM5* for Pearson’s correlation coefficients).

**Figure 5:**
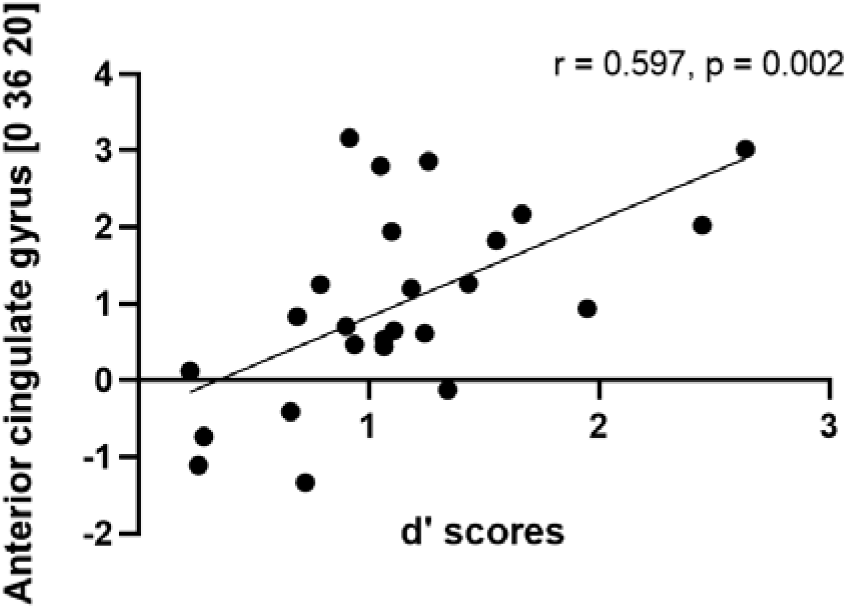
Association of task-induced activation in anterior cingulate gyrus with (d’) scores.

No significant association was found between age or socio-economic status, or BRIEF scores and cerebral activations related to response inhibition for both tasks.

## 4 Discussion

### Sustained attention performance using the gradCPT paradigm

Our behavioral results showed that the *scene* gradCPT was better performed compared to the *face* gradCPT. This was evidenced by higher sensitivity scores (*d’*) and lower reaction time variability (RT_CV_).

Indeed, higher *d’* scores suggest that participants were more sensitive and demonstrated better discrimination processes during the *scene* gradCPT rather than during the *face* gradCPT. We can hypothesize that cities and mountains stimuli are more discernible compared to male and female faces, leading to a stronger bottom-up attention that improves the ability to differentiate between stimuli.

Regarding the RT_CV_, lower mean values of RT variability, reflecting better sustained attention (Manly et al., 2000; O’Halloran et al., 2018), were observed in the *scene* compared to the *face* gradCPT. In addition, Esterman et al. (2013) considered the possibility that low variability could indicate states when motor response settings are optimized to balance between speed and accuracy. Therefore, participants were more accurate and able to better adjust their motor responses for cities and mountains stimuli during the *scene* gradCPT. Furthermore, RT_CV_ might be influenced by factors such as the complexity of the modality for instance. We can argue that higher variability of responses can be due to a more cognitively challenging task for participants and that the *face* gradCPT stimuli have higher degree of similarities than the *scene* gradCPT stimuli.

Accuracy analyses revealed a significant difference between the percentages of correct NT and correct T in both tasks, meaning that participants could respond precisely to NT stimuli, while they faced difficulties in inhibiting their motor responses for T stimuli. In the current study, we observed lower percentages of correct responses to T stimuli (*face* gradCPT: percentage of correct T = 22.1%; *scene* gradCPT: percentage of correct T = 28.1%) compared to the previous studies conducted in adults population and using the same gradCPT paradigm (percentage of correct T = 74-80%) (Esterman et al., 2013; Rosenberg et al., 2013). This discrepancy may be attributable to a less mature inhibition system and increased impulsive reactions during adolescence. Indeed, the presence of impulsive behaviors in adolescents is well-documented and has been already studied (Fortenbaugh et al., 2015; Halperin et al., 1991). This hypothesis of impulsivity is also supported by shorter mean reaction times observed in the current study (*face* gradCPT: M = 569ms; *scene* gradCPT: M = 577ms) compared to those reported in previous studies conducted in adult participants (M = 906 ms, Rosenberg et al., 2013).

Using RT_CV_, we found two significant effects on behavioral performances of covariates of interest, including the number of errors in the Go/No-Go task (2 stimuli, 1 target) and the Conners inattention scores. Less impulsive participants (those making less errors in the Go/No-Go task) paradoxically showed more variability in RT responses and lower performance during the gradCPT paradigm. We could argue that this is a strategic approach adopted by participants to balance between accuracy and speed during their performance. Less impulsive participants might be more accurate by responding slower, decreasing their error rates but demonstrating higher variability in RTs. On the opposite, more impulsive participants might respond faster, which leads to a higher error rate but interestingly more consistent RTs. These findings may appear not intuitive with expectations and call for further explorations.

In addition, we observed that inattentive participants showed higher RT_CV_ scores (more variability in responses, lower performance). This aligns with a previous study that associated greater variability in RT with increased inattention in ADHD individuals (Adams et al., 2011). Our results showed that factors such as number of errors and the inattention scores might be predictors of adolescents’ behavioral performances during the gradCPT paradigm.

### Whole-brain activations of response inhibition during the gradCPT paradigm

Investigating the main effect of response inhibition regardless of the type of modality (T > NT), our findings revealed a bilateral fronto-parieto-occipito pattern of brain acitivity, that overlaps with the sustained attention network described by Langner & Eickhoff (2012) in adults and by Morandini et al. (2020) in young adolescents. We observed a strong frontal activity in many regions involved in executive functions and planning brain regions but also in motor control. Indeed, the ACC has been involved in conflict monitoring and plays an important role in the detection of errors and response adjustments (Bediou et al., 2012; Ridderinkhof et al., 2004). This region is known to be highly connected to and works closely with the anterior insula, a brain region involved in detecting relevant stimuli and facilitating the processing of task-related information (Menon & D’Esposito, 2022).

Moreover, parietal regions also seem to be recruited for response inhibition. Activity in the SPL and the precuneus are known to allow a top-down (voluntary) control of attention and an attention shifting (Koenigs et al., 2009; Yantis et al., 2002; Yantis & Serences, 2003). The IPS has been involved in attentional readjustments in case of attentional drifts and enables redirection toward the task and an efficient maintain of sustained attention (Corbetta et al., 2008; Langner & Eickhoff, 2012; Malhotra et al., 2009; Posner M, Petersen J., 1990).

Finally, we observed activity in occipital areas that support the processing and identification of stimuli. The fusiform gyrus is a well-known structure for high-level visual processes such as facial and object recognition (Weiner & Zilles, 2016). Moreover, the observed bilateral activation of the posterior cerebellum also corroborate previous studies showing its involvement in cognitive attention processes and its contribution in sustained attention performance (Buckner, 2013; Langner & Eickhoff, 2012; Michael et al., 2009; Stoodley, 2012).

In addition to cortical activations, thalamic activity has been found in the current study as well as in previous sustained attentional study (Langner & Eickhoff, 2012). These authors suggested that the thalamus may be implicated in maintaining arousal during the task. Another hypothesis made by these authors is the implication of the thalamus in a supplementary or compensatory attentional effort.

Overall, our investigation of the main effect of response inhibition revealed bilateral brain activations that includes frontal, parietal, occipital, and thalamic brain regions. These results suggest that adolescents seem to recruit brain areas typically involved in the performance of a sustained attention task such as the gradCPT paradigm.

### Whole-brain activations during the face and scene gradCPT

The main effect of response inhibition (T > NT) during the *face* gradCPT showed a mainly left-lateralized pattern of activation in fronto-cingulo-cerebellum regions. An extended bilateral pattern of fronto-temporo-parieto-occipital activity was recruited during the *scene* gradCPT, with additional brain regions such as the anterior cingulate cortex, the anterior insula, the cerebellum, and the thalamus. As previously described, the anterior cingulate cortex and the anterior insula are key brain regions involved in completion of sustained attention tasks and were already associated with a good performance in healthy and ADHD adolescents during a go/no-go task (O’Halloran et al., 2018). Such brain activations observed only during the *scene* gradCPT align with our previous discussion regarding the better performance in the *scene* gradCPT, as well as the distinction of *scene* compared to *face* stimuli, where city and mountain stimuli were notably more distinguishable than those involving male and female faces.

### Influence of covariates of interest on brain activations related to response inhibition

In the current study, we observed positive significant correlations between brain activity and gradCPT behavioral attentional performances (for both *d’* scores and RT_CV_), Conners 3-SR Inattention scores, and TAP scores, specifically in the Go/No-Go tasks.

Regarding the significant correlations with gradCPT behavioral attentional performances, adolescents with higher *d’* scores displayed stronger brain activity in the left ACC during response inhibition in the *scene* gradCPT specifically. Although we observed a correlation specifically in the left ACC, our results are in line with the study of O’Halloran et al. (2018), who found an association between increased bilateral activation in the ACC and increased performance during sustained attention task. As discussed previously, left brain regions activation seemed to reflect the challenging nature of an attentional task and high cognitive demands. Furthermore, adolescents with higher RT_CV_ (higher variability in RT responses, meaning pooper performance) showed increased brain activity in frontal, parietal, and temporal areas during response inhibition, specifically in the *face* gradCPT. These findings are consistent with previous studies linking higher behavioral variability in response to the inhibition task with increased activity in fronto-parietal areas (Bellgrove et al., 2004; Simmonds et al., 2007). Regarding the *scene* gradCPT, higher variability in RT responses was associated with higher brain activity in the left posterior cerebellum. A prior study using Go/No-Go tasks also associated higher variability but in the right posterior cerebellum (Simmonds et al., 2007). Our study is the first demonstrated positive correlations between RT variability and the left posterior cerebellum and needs confirmation and further investigations.

Our investigations of effects of covariates of interests on sustained attention cerebral activation revealed that participants with higher levels of inattention, as measured by a self-reported questionnaire, displayed increased activity in frontal, temporal, parietal, occipital brain regions, and the right posterior insula during the *face* gradCPT. During the *scene* gradCPT, we observed that higher inattention scores were associated with increased brain activity in parietal and occipital regions, and in the left posterior cerebellum. We can thus hypothesize that more inattentive adolescents might use compensatory mechanisms to perform the task similarly to the less inattentive ones, as we observed an increased activity in brain regions typically involved in sustained attention processes.

Finally, we found that brain activity during the *scene* and the *face* gradCPT were positively associated with the number of errors during Go/No-go computerized tasks from the TAP. In other words, adolescents making more errors during the Go/No-Go task from the TAP showed higher impulsive responses (that may indicate inhibitory control deficit) that correlated with stronger brain activity in specific regions during the gradCPT tasks. We thus demonstrated a link between impulsive behaviour and neural activity related to response inhibition. In addition, Menon et al (2001) studied the error-related brain activity linked with failure to inhibit response during a Go/No-Go task and demonstrated that brain regions involved in error processing partially overlap with those involved in response inhibition. Based on these studies and on our findings, the following hypothesis could be proposed: adolescents, due to impulsive reactions, struggled to inhibit their dominant responses and were conscious of the errors they made. Consequently, brain regions associated with error processing were more prominently engaged in response to these mistakes.

## 5 Limitations

In contrasts to Rosenberg et al. (2013) findings in adults, our study encountered challenges with adolescent participants. The percentage of correct target trials were lower for both modalities (*scene* and *face*), meaning that adolescents struggled in inhibiting responses. Thus, rather than selecting only the correct target trials for whole-brain activation investigations, we choose to include all target trials.

One potential reason for poor task performance may be related to the complexity of the task itself. We previously mentioned that adolescents might answer in a more impulsive way and might have difficulties in correctly withholding to target stimuli specifically. However, we do not believe that participants were disinterested by the gradCPT and responded in an arbitrary manner. If participants had responded randomly, the error rate in the nontarget condition (omission errors) would have been close to 50%, which is greater than ours. In addition, a lack of interest in the tasks would not have induced brain activity associated with response inhibition. Besides, our findings on neural substrates of sustained attention and inhibition are in line with those found in previous studies. Furthermore, our behavioral performances scores (*d’* specifically) during the *scene* gradCPT are close to those found in previous studies in adults (M= 1.18, Rosenberg et al., 2013). Regarding the *face* gradCPT specifically, there are no comparable studies available. We recognize that behavioral performances scores in the *face* gradCPT were lower than expected and task-induced brain activity seemed to be weaker compared to the *scene* gradCPT.

Finally, we can also discuss the gradCPT paradigm applied in an adolescent population. Although the gradCPT paradigm has been validated as a valuable tool to assess sustained attention and inhibition, its application on children or adolescent populations is still limited. A recent study investigated sustained attention performance across the life span in 10’000 participants (Mean age = 26.07, SD = 11.77, range : 10-70 years) using the gradCPT test and demonstrated an increase in performances between 10 to 16 years-old, with peak ability in sustaining attention around the mid-40s (Fortenbaugh et al., 2015). McAvinue et al. (2012) found similar findings using the sustained-attention-to-response task and showed that performance varied, being weaker during childhood and adolescence, stabilizing in young and middle adulthood, and finally declining in older adulthood. Thus, these findings might also explain why behavioral performances during the *scene* gradCPT were slightly lower than those in adults, although adolescents seemed to recruit brain areas typically involved in the performance of a sustained attention task.

In sum, this study provides a better comprehension of the neural substrates associated with sustained attention and inhibition processes in typically developing adolescents using the gradCPT paradigm. More specifically and consistent with previous studies, we observed brain activity related to sustained attention in bilateral fronto-parieto-occipito brain regions.

## Data Availability

All data produced in the present study are available upon reasonable request to the authors

## Acknowledgments

We thank and acknowledge all participating adolescents and their families who made this research possible. We also thank the Foundation Campus Biotech Geneva (FCBG), a Foundation of the Ecole Polytechnique Fédérale de Lausanne (EPFL), the University of Geneva, and the University Hospitals of Geneva. Finally, we extend our gratitude to the pediatric clinical research platform of the Geneva University Hospital for their help in data management.

