## Supplemental Material for "Assessing sustained attention processes and related cerebral activations in typically developing adolescents using the gradual-onset continuous performance task (gradCPT)"

*Shared authorship

### Classification of the target and non-trial trials

Based on Rosenberg and colleagues (2013), an accurate non-target trial happened when participants pressed the response button within a time range of 560ms and 1’520ms after the stimulus onset. Any ambiguous presses (highly deviant reaction times, meaning shorter than 560ms or longer than 1’520ms) or multiple button presses during a single trial were assessed using an iterative algorithm from Rosenberg et al. (2013) to maximize correct responses and potentially assign to adjacent trials. Finally, an accurate target trial happened when participants withheld their response between 560ms and 1’520ms after the stimulus onset.

***Table SM1:*** *Regional cerebral activations during the Face and Scene tasks for the main effect of tasks. (A) Common brain regions activated in both Face and Scene tasks (Conjunction analysis). (B) and (C) Differences in brain activations between the Face and Scene tasks. All reported clusters are significant at peak-level at p < 0.001, uncorrected for multiple comparisons, with a minimum size of 50 voxels. Peaks that are significant after correction for multiple comparison are indicated with an asterisk (*FDR-corrected).*

*Abbreviation: Lat.: Hemisphere lateralisation (L = left; R = right), Z-score ($Z_E$) values refer to the activation maxima to the SPM coordinates.*

| Cerebral regions | | Lat. | MNI coordinates x | y | z | Cluster size | $Z_E$ |
|---|---|---|---|---|---|---|---|
| (A) Face & Scene | | | | | | | |
| *Frontal* | Posterior-medial frontal – dACC/SMA | L | 0 | 18 | 44 | 155 | 3.995 |
| | Posterior-medial frontal – dACC/SMA | R | 16 | 10 | 62 | 183 | 3.761 |
| | Superior – precentral gyrus | L | -26 | -2 | 60 | 118 | 3.394 |
| | Superior frontal gyrus | L | -22 | 0 | 52 | 118 | 4.267 |
| | Superior frontal gyrus | R | 26 | 4 | 64 | 183 | 3.325 |
| *Other* | Cerebellum (VI) | L | -40 | -46 | -30 | 112 | 4.254 |
| (B) Face > Scene | | | | | | | |
| *Temporal* | Middle temporal gyrus | R | 44 | -58 | 4 | 73* | 4.584 |
| (C) Scene > Face | | | | | | | |
| *Temporal* | Superior parahippocampal gyrus | L | -28 | -48 | -8 | 2706* | 7.325 |
| | Superior parahippocampal gyrus | R | 22 | -38 | -16 | 3326* | 7.159 |
| *Parietal* | Inferior precuneus | L | -18 | -56 | 14 | 100 | 4.337 |
| | Inferior precuneus | R | 18 | -50 | 22 | 273 | 3.903 |
| | Superior parietal lobule | R | 18 | -78 | 48 | 109 | 4.204 |
| *Occipital* | Calcarine gyrus | R | 16 | -94 | -4 | 3326* | 4.472 |
| | Posterior cuneus | R | 16 | -98 | 6 | 3326 | 4.212 |
| | Inferior occipital gyrus | L | -28 | -78 | -2 | 2706* | 4.499 |
| | Superior lingual gyrus | R | 26 | -60 | -8 | 3326* | 6.749 |
| | Middle occipital gyrus | L | -24 | -80 | 20 | 2706* | 5.497 |
| | Middle occipital gyrus | R | 32 | -76 | 16 | 3326* | 6.013 |
| *Other* | Cingulate cortex - Middle cingulate gyrus | L | -8 | 14 | 38 | 155 | 3.224 |

***Table SM2:*** *Regional cerebral activations during response inhibition (Target > Non-Target) for the interaction Tasks x Stimuli. All reported clusters are significant at peak-level at p < 0.001, uncorrected for multiple comparisons, with a minimum size of 50 voxels. Peaks that are significant after correction for multiple comparison are indicated with an asterisk (*FDR-corrected).*

*Abbreviation: Lat.: Hemisphere lateralisation (L = left ; R = right), Z-score ($Z_E$) values refer to the activation maxima to the SPM coordinates.*

| Cerebral regions | | | MNI coordinates | | | | |
|---|---|---|---|---|---|---|---|
| | | **Lat.** | **x** | **y** | **z** | **Cluster size** | **$Z_E$** |
| Scene (T>NT) > Face (T>NT) | | | | | | | |
| *Frontal* | Inferior frontal gyrus (*pars opercularis*) | R | 48 | 16 | 0 | 397 | 3.424 |
| | Middle frontal gyrus | R | 42 | 28 | 36 | 385 | 3.601 |
| | Inferior precentral gyrus | R | 50 | 8 | 34 | 385 | 3.764 |
| *Parietal* | Intraparietal sulcus | R | 40 | -42 | 46 | 587 | 3.776 |
| | Inferior parietal lobule | R | 48 | -36 | 52 | 587 | 4.255 |
| | Superior parietal lobule | L | -22 | -64 | 34 | 53 | 3.524 |
| *Occipital* | Superior fusiform gyrus – FFA | R | 32 | -62 | -12 | 101 | 4.144 |
| | Inferior fusiform gyrus (area FG3) | L | -30 | -58 | -14 | 282 | 4.047 |
| | Inferior fusiform gyrus (area FG3) | R | 28 | -48 | -12 | 70 | 3.490 |
| | Inferior lingual gyrus | L | -22 | -46 | -10 | 282 | 3.246 |
| | Middle occipital gyrus | L | -26 | -78 | 22 | 128 | 4.025 |
| *Other* | Anterior insula | L | -34 | 18 | -2 | 294 | 4.719 |
| | Anterior insula | R | 28 | 18 | -8 | 397* | 5.351 |
| | Cerebellum (crus 1) | L | -40 | -58 | -28 | 282 | 3.698 |
| | Cerebellum (IV-V) | R | 30 | -42 | -22 | 70 | 3.599 |
| | Cerebellum (VI) | L | -32 | -56 | -32 | 282 | 3.471 |
| | Thalamus | R | 10 | -10 | 0 | 135 | 4.175 |

***Table SM3***: *Pearson's correlations between individual Beta values and d' and $RT_{CV}$ scores during the GradCPT tasks. (*) significant p-value.*

| | **Cerebral regions** | **Lat.** | **Pearson's correlation coefficient (r)** | **p-value** |
|---|---|---|---|---|
| Face task | **Association with $RT_{CV}$** | | | |
| *Frontal* | Superior frontal gyrus | L | .589 | .0002* |
| | Superior frontal gyrus | R | .709 | <.0001* |
| | Posterior-medial frontal – dACC/SMA | R | .633 | .001* |
| *Temporal* | Middle temporal gyrus | L | .655 | .0004* |
| | Superior temporal gyrus | L | .697 | .0001* |
| *Parietal* | Angular gyrus | L | .673 | .0002* |
| | Angular gyrus | R | .710 | <.0001* |
| Scene task | **Association d' scores** | | | |
| *Other* | Anterior cingulate cortex | L | .597 | .002* |
| | **Association with $RT_{CV}$** | | | |
| *Other* | Cerebellum (VI) | L | .396 | .050* |

***Table SM4:*** *Pearson's correlations between individual Beta values and Conners' inattention scores during the GradCPT tasks. (*) significant p-value.*

| | Cerebral regions | Lat. | Pearson's correlation coefficient (r) | p-value |
|---|---|---|---|---|
| Face task | **Association with Conners' inattention scores** | | | |
| *Frontal* | Superior precentral gyrus | L | 0.337 | 0.100 |
| *Temporal* | Superior temporal gyrus | L | 0.319 | 0.120 |
| *Parietal* | Superior precuneus | L | 0.444 | 0.026* |
| | Superior parietal lobule | L | 0.412 | 0.041* |
| | Superior parietal lobule | R | 0.504 | 0.010* |
| *Occipital* | Calcarine sulcus | L | 0.752 | <.001* |
| | Calcarine sulcus | R | 0.584 | 0.002* |
| | Superior cuneus | L | 0.465 | 0.019* |
| | Superior cuneus | R | 0.551 | 0.004* |
| | Lingual gyrus | L | 0.494 | 0.012* |
| | Superior occipital gyrus | L | 0.498 | 0.011* |
| | Superior occipital gyrus | R | 0.425 | 0.034* |
| *Other* | Insula lobe – posterior insula | R | 0.469 | 0.018* |
| | Parahippocampal gyrus | L | 0.602 | 0.002* |
| Scene task | | | | |
| *Parietal* | Precuneus | L | 0.401 | 0.047* |
| *Occipital* | Calcarine sulcus | L | 0.316 | 0.124 |
| | Cuneus | L | 0.149 | 0.479 |
| | Cuneus | R | 0.343 | 0.094 |
| | Inferior lingual gyrus | L | 0.342 | 0.095 |
| | Inferior lingual gyrus | R | 0.205 | 0.325 |
| | Superior occipital gyrus | L | 0.286 | 0.162 |
| | Superior occipital gyrus | R | 0.415 | 0.039* |
| *Other* | Cerebellum (VI) | L | 0.333 | 0.104 |

***Table SM5****: Pearson's correlations between individual Beta values and TAP scores during the GradCPT tasks. (*) significant p-value.*

| | Cerebral regions | Lat. | Pearson's correlation coefficient (r) | p-value |
|---|---|---|---|---|
| Face task | **Association with Go/No-go scores (2 stim – 1 target)** | | | |
| *Parietal* | Inferior precuneus | L | 0.655 | <0.001* |
| *Occipital* | Inferior cuneus | L | 0.694 | <0.001* |
| *Subcortical* | Basal ganglia – putamen | L | 0.704 | <0.001* |
| | **Association with Go/No-go scores (5 stim – 2 targets)** | | | |
| *Frontal* | Superior precentral gyrus | L | 0.661 | <0.001* |
| Scene task | **Association with Go/No-go scores (5 stim – 2 targets)** | | | |
| *Temporal* | Superior temporal gyrus | R | 0.752 | <0.001* |
| *Other* | Insula lobe – posterior insula | R | 0.656 | <0.001* |
